# Atherogenic index of plasma is associated with the risk of myocardial infarction: a prospective cohort study

**DOI:** 10.1101/2023.02.16.23286068

**Authors:** Yijun Zhang, Shouling Wu, Xue Tian, Qin Xu, Xue Xia, Xiaoli Zhang, Jing Li, Shuohua Chen, Anxin Wang, Fen Liu

## Abstract

**Background and aims:** The atherogenic index of plasma (AIP) has been confirmed as a contributor of cardiovascular disease. But few evidence on the longitudinal pattern of AIP during follow-up. This study aimed to explore the associations between baseline and long-term AIP with the risk of myocardial infarction (MI).

**Methods:** A total of 98 861 participants without MI at baseline were included from the Kailuan study. The baseline AIP was calculated as log (triglyceride/high-density lipoprotein cholesterol). The long-term AIP was calculated as the updated mean AIP and the number of visits with high AIP. The updated mean AIP was calculated as the mean of AIP from baseline to the first occurrence of MI or to the end of follow-up. The number of visits with high AIP was defined as higher than the cutoff value at the first three visits. Univariable and multivariable Cox proportional hazard models were used to determine the association between AIP and the risk of MI.

**Results:** During a median follow-up of 12.80 years, 1804 participants developed MI. The multivariable models revealed that elevated levels of baseline and updated mean AIP increased the risk of MI, compared with quartile 1 the HR in quartile 4 was 1.63 (95% CI, 1.41-1.88) and 1.59 (95% CI, 1.37-1.83), respectively. Compared to those without high AIP, the risk of individuals with three times was 1.94 (95% CI,1.55-2.45).

**Conclusions:** Elevated levels of both baseline and long-term AIP displayed a higher risk of MI.

## Introduction

Cardiovascular disease (CVD) is the leading cause of global mortality and a major contributor to disability, and myocardial infarction (MI) served as the main causes of CVD^1^. CVD mortality has declined in high-income areas, but still seriously in developing countries^2^. Thus, it’s necessary to early identify population at high-risk of MI.

Disorders of lipoprotein metabolism named dyslipidemia are thought to be the one of most important risk factors for MI^3^. Elevated level of total cholesterol (TC), triglyceride (TG), low-density lipoprotein cholesterol (LDL-C), and the high ratio of LDL-C to high-density lipoprotein cholesterol (HDL-C) increased the risk of MI^4-6^. The atherogenic index of plasma (AIP) is calculated as log (TG/HDL) and reflects the levels of TG and HDL-C cholesterol, which is considered a potential biomarker of MI^7-10^. It was reported that AIP was a potential biomarker in the early diagnosis of MI from a cross-sectional study^8^. A hospital-based observational study suggested that AIP could predict the size of lipoprotein particles, showing a positive correlation with the risk of MI^9^. Additionally, elevated AIP was reported as a powerful independent predictor of MI^11-14^ and arterial stiffness^15^ beyond traditional risk factors^16^. The previous studies always focused on specific individuals, or based on a small sample, lacked of the evidence based on the large sample in general individual.

Most previous studies focused on the effect of AIP on clinical outcomes in MI had used one time-point AIP. This approach has a limited power to predict the outcome as changes of many biological and environmental factors during long-term follow-up could impact the level of lipid, a single measurement of AIP may lead to incorrect classification of risk assessment for MI. Instead, the updated AIP measured at multiple time points can characterize the longitudinal pattern of AIP, thus providing a more robust assessment of associations with outcomes than single measurement of AIP. Therefore, the purpose of our study was to explore whether elevated AIP at baseline and in the long-term, which the long-term AIP was defined as updated mean of AIP and number of visits with high AIP, were associated with a higher risk for MI based on a large community prospective cohort study.

## Methods

### Study population

The Kailuan Study is an ongoing prospective cohort study that was performed in the community of Kailuan in Tangshan, China^17^. This study enrolled 101 510 individuals (81 110 men and 20 400 women) with an age ranging from 18 to 98 years at baseline during June 2006 and October 2007. All the participants provided written informed consent, and biennially carried out questionnaire assessments, clinical examinations, and laboratory tests. The details of this study design have been published previously^18^. The study was performed according to the guidelines of the Helsinki Declaration and was approved by the Ethics Committee of Kailuan Hospital (approval number: 2006e05) and Beijing Tiantan Hospital (approval number: 2010-014-01).

In this present study, we analyzed the associations between baseline (2006) and long-term AIP with the risk of MI. Data analysis was performed from 1 January 2006 to 31 December 2019. We included 98 861 participants free of MI and without missing data of TG and HDL-C at baseline for the analysis of the baseline and updated mean AIP. We further excluded 42 654 participants with missing data of TG and HDL-C and 195 with developed MI during 2006 and 2010 for the analysis of number of visits with high AIP. The flowchart of this study cohort is presented in Figure 1.

**Figure. 1.**
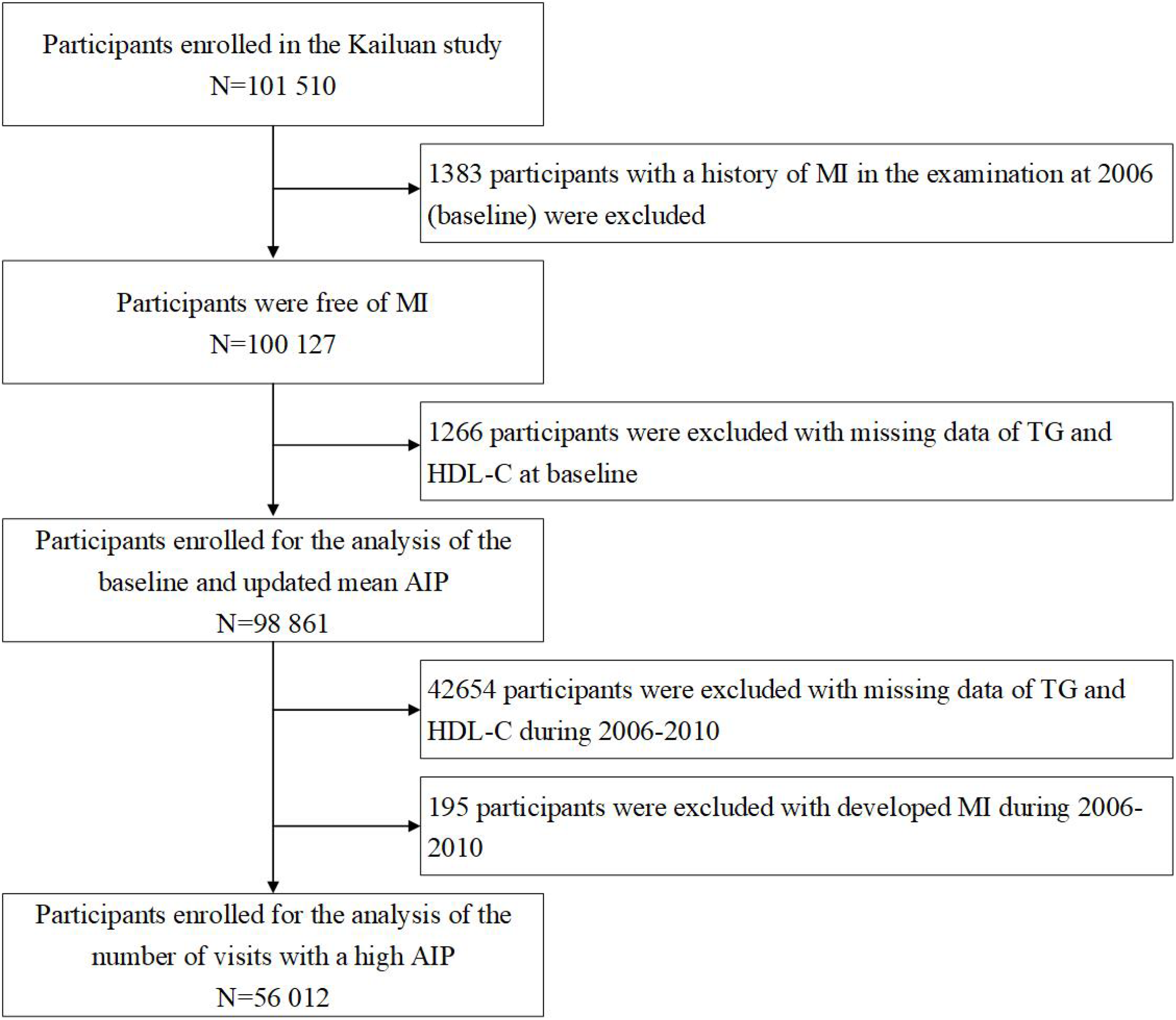
The flowchart of the study. Abbreviation: MI, myocardial infarction; TG, triglyceride; HDL-C, high-density lipoprotein cholesterol; AIP, atherogenic index of plasma.

### Data collection and definitions

Demographic information (such as age, sex, educational levels, and income), lifestyle (such as current smoking, drinking, and physical exercise habits), and disease history (such as hypertension, hyperlipidemia, and diabetes) were obtained through questionnaires. Educational levels were classified as illiteracy or primary school, middle school, and high school or above. Income was classified as >800 and ≤800 yuan per month. Smoking and drinking status were classified as yes or no. Active physical exercise was defined as ≥4 times per week and ≥20 min at a time. Blood pressure was measured in the seated position using a mercury sphygmomanometer, and the average of three readings was calculated as systolic blood pressure (SBP) and diastolic blood pressure (DBP). Blood was drawn from the antecubital vein in the morning after 12 hours fast for determinations of fasting plasma glucose (FPG), serum TC, TG, LDL-C, and high-density lipoprotein cholesterol (HDL-C). All the plasma samples were assessed using an auto-analyzer (Hitachi 747, Tokyo, Japan) at the central laboratory of Kailuan Hospital. FPG levels were measured using the hexokinase/glucose-6-phosphate dehydrogenase method with the coefficient of variation using blind quality control specimens <2.0%. TC, TG, LDL-C and HDL-C levels were measured with the enzymatic colorimetric method.

### Calculation of the baseline and long-term AIP

The AIP was a logarithmically transformed ratio of TG to HDL-C in molar concentration (mmol/L), and it was mathematically derived from log (TG/HDL-C)^7^. To determine long-term AIP patterns of individuals, we calculated the updated mean AIP and the number of visits with high AIP. The updated mean AIP was calculated using the average value of the AIP from baseline to the date of onset of MI, death, or to the end of follow-up (2019), whichever came first. Hence, those participants who did not develop MI before the end of follow-up had seven times of AIP measurements and those who developed MI had less than seven times.

In this study, high AIP was defined as higher than the cutoff value which was estimated by the receiver operating characteristic curve (ROC). And the number of visits with high AIP was using the AIP at the 1^st^, 2^nd^ and 3^rd^ visit, which was assigned 1 point to higher than the cutoff value of AIP and 0 point to lower at each visit, so the number of visits with a high AIP was ranged from 0 to 3, such as 2 points expressed with two times high in all the three visits.

### Follow-up and assessment of MI

Follow-up ended at the date of onset of MI, death, or to the end of follow-up (2019), whichever came first. The primary outcome was the first occurrence of MI, either the first nonfatal MI event or death due to MI without a history of MI. We used 10th Revision code I21 for MI. The database of MI diagnoses was obtained from the Municipal Social Insurance Institution and Hospital Discharge Register from the 11 hospitals and was updated annually during the following period. The diagnosis of MI was obtained from the patient’s clinical symptoms, electrocardiogram, and dynamic changes of myocardial enzyme following the World Health Organization’s Multinational Monitoring of Trends and Determinants in Cardiovascular Disease criteria, and the criteria were consistently applied across all 11 hospitals^19^.

### Statistical analysis

The population was divided into four groups by baseline or updated mean AIP quartiles, and the number of visits with a high AIP. If the continuous variables were normally distributed, then they were described by mean and standard deviation, and the group differences were compared by one-way analysis of variance (ANOVA). Otherwise, the continuous variables were described by median and interquartile range (IQR), and the group differences were compared by Kruskal-Wallis tests. The categorical variables were described by frequency (percentages), and were compared by chi-square tests to analyze group differences. Person-years at risk was computed from the date of baseline until the date of ending follow-up in this study or the date of onset of MI, death, whichever came first.

Cox proportional hazards regression models were used to estimate the association of AIP with MI incidence risk by calculating the hazard ratio (HR) and 95% confidence interval (CI). There were three cox proportional hazards regression models: Model 1 was unadjusted; Model 2 was adjusted for age and sex at baseline; Model 3 was further adjusted for education, smoking status, drinking status, body mass index (BMI), SBP, FPG, high sensitivity C-reactive protein (hs-CRP), TC, and LDL-C level, history of hypertension, hyperlipidemia, diabetes, antihypertensive drugs, antidiabetic drugs, lipid-lowering drugs at baseline. The *P* values for trend were computed using quartiles of baseline or the updated mean AIP as ordinal variables, and using restricted cubic spline (RCS) with 5 knots (at the 5th, 25th, 50th, 75th, and 95th percentiles) to analyze the effect of baseline and the updated mean AIP on MI as a continuous variable. The ROC curve analysis was used to determine the best cutoff value of AIP in case of incident MI. The best cutoff point of AIP was assessed by the maximum value of the Youden index, which was calculated as sensitivity+specificity−1.

To evaluate the robustness of the association of AIP with the risk of MI, sensitivity analyses were applied, adjusted for variables in model 3 and further excluded all deaths during the follow-up visits. Stratified analyses according to baseline age (≤65 vs. >65 years), sex, BMI (< 24 vs. ≥24 kg/m^2^), hyperlipidemia (no vs. yes), and lipid-lowering drugs (no vs. yes) were used to examine the consistence of the effect of baseline AIP (quartile 4 group) for the risk of MI.

All *P* values were two-sided, with *P*< 0.05 considered statistically significant. All statistical analyses were performed using SAS Version 9.4 software (SAS Institute, Cary, NC, USA).

## Results

This study included 98 861 participants with the median age was 51.81 years (IQR, 43.60 to 59.23) and 78 849 (79.76%) were men. The average baseline AIP level was - 0.16 (IQR, -0.56 to 0.31). Table 1 showed baseline characteristics by quartiles of baseline AIP. In general, individuals with higher AIP were more likely to be older, men, lower educational, less income, inactive physical activity, more current smokers and drinkers, a higher prevalence of hypertension, diabetes, and dyslipidemia, more likely to take antihypertensive drugs, antidiabetic drugs, and lipid-lowering drugs, and had higher BMI, SBP, FPG, hs-CRP, TC and LDL-C levels. The group differences between the quartiles of the updated mean AIP produced the same result (Table S1).

**Table 1.** Baseline characteristics of participants according to quartiles of baseline AIP

| Characteristics | Overall | Quartiles of baseline AIP |  |  |  | *P* |
| --- | --- | --- | --- | --- | --- | --- |
|  |  | Q1 | Q2 | Q3 | Q4 |  |
| No. of participants | 98861 | 24717 | 24718 | 24709 | 24717 |  |
| Baseline AIP | -0.16 (-0.56-0.31) | -0.84 (-1.06--0.68) | -0.35 (-0.45--0.26) | 0.05 (-0.06-0.17) | 0.70 (0.48-1.02) | <0.0001 |
| Age, year | 51.81 (43.60-59.23) | 51.65 (43.20-59.80) | 51.72 (43.46-59.31) | 52.29 (44.09-59.53) | 51.55 (43.63-58.35) | <0.0001 |
| Men, n (%) | 78 849 (79.76) | 18 431 (74.57) | 19 469 (78.76) | 20 013 (80.99) | 20 936 (84.70) | <0.0001 |
| High school or above, n (%) | 19 187 (19.41) | 5424 (21.94) | 4361 (17.64) | 4629 (18.73) | 4773 (19.31) | <0.0001 |
| Income >800 RMB/month, n (%) | 13 661 (13.82) | 3665 (14.83) | 3106 (12.57) | 3363 (13.61) | 3527 (14.27) | <0.0001 |
| Active physical activity, n (%) | 87 013 (88.02) | 21 859 (88.44) | 21 966 (88.87) | 21 696 (87.81) | 21 492 (86.95) | <0.0001 |
| Current smoking, n (%) | 33 071 (33.45) | 7903 (31.97) | 7531 (30.47) | 8309 (33.63) | 9328 (37.74) | <0.0001 |
| Current drinking, n (%) | 35 951 (36.37) | 9116 (36.88) | 8064 (32.62) | 8824 (35.71) | 9947 (40.24) | <0.0001 |
| BMI, kg/m<sup>2</sup> | 24.84 (22.60-27.22) | 23.18 (21.19-25.39) | 24.45 (22.41-26.67) | 25.40 (23.35-27.66) | 26.30 (24.22-28.41) | <0.0001 |
| SBP, mmHg | 130.00 (119.30-141.30) | 121.30 (110.70-140.00) | 129.30 (117.30-140) | 130.00 (120.00-143.30) | 130.00 (120.00-149.30) | <0.0001 |
| Hypertension, n (%) | 12 307 (12.45) | 2019 (8.17) | 2608 (10.55) | 3520 (14.25) | 4160 (16.83) | <0.0001 |
| Hyperlipidemia, n (%) | 3033 (3.07) | 493 (1.99) | 605 (2.45) | 864 (3.50) | 1071 (4.33) | <0.0001 |
| Diabetes Mellitus, n (%) | 5758 (5.82) | 857 (3.47) | 1096 (4.43) | 1563 (6.33) | 2242 (9.07) | <0.0001 |
| Antihypertensive drugs, n (%) | 10 675 (10.80) | 1708 (6.91) | 2281 (9.23) | 3037 (12.29) | 3649 (14.76) | <0.0001 |
| Antidiabetic drugs, n (%) | 2311 (2.34) | 376 (1.52) | 462 (1.87) | 662 (2.68) | 811 (3.28) | <0.0001 |
| Lipid-lowering drugs, n (%) | 846 (0.86) | 121 (0.49) | 156 (0.63) | 215 (0.87) | 354 (1.43) | <0.0001 |
| FPG, mmol/L | 5.11 (4.66-5.71) | 4.99 (4.53-5.48) | 5.10 (4.65-5.62) | 5.19 (4.70-5.80) | 5.29 (4.77-6.01) | <0.0001 |
| hs-CRP, mg/dL | 0.80 (0.30-2.20) | 0.60 (0.21-1.90) | 0.74 (0.29-2.00) | 0.90 (0.34-2.30) | 1.01 (0.40-2.60) | <0.0001 |
| TC, mg/dL | 4.92 (4.28-5.59) | 4.79 (4.20-5.41) | 4.91 (4.28-5.51) | 5.01 (4.39-5.67) | 5.01 (4.26-5.75) | <0.0001 |
| LDL-C, mg/dL | 2.33 (1.82-2.83) | 2.18 (1.64-2.74) | 2.40 (1.95-2.84) | 2.40 (1.94-2.90) | 2.31 (1.77-2.83) | <0.0001 |

Abbreviation: AIP, atherogenic index of plasma; BMI, body mass index; SBP, systolic blood pressure; FPG, fasting plasma glucose; hs-CRP, high-sensitivity C-reactive protein; TC, total cholesterol; LDL-C, low-density lipoprotein cholesterol.

During a median follow-up of 12.80 years (IQR, 12.37 to 13.30), 1804 (1.82%) participants developed MI. As shown in Table 2, with the higher baseline AIP level, the incidence of MI was higher, from 0.96 (95% CI, 0.86-1.08) in quartile 1 to 1.91 (95% CI, 1.76-2.07) per 1000 person-years in quartile 4.

**Table 2.**
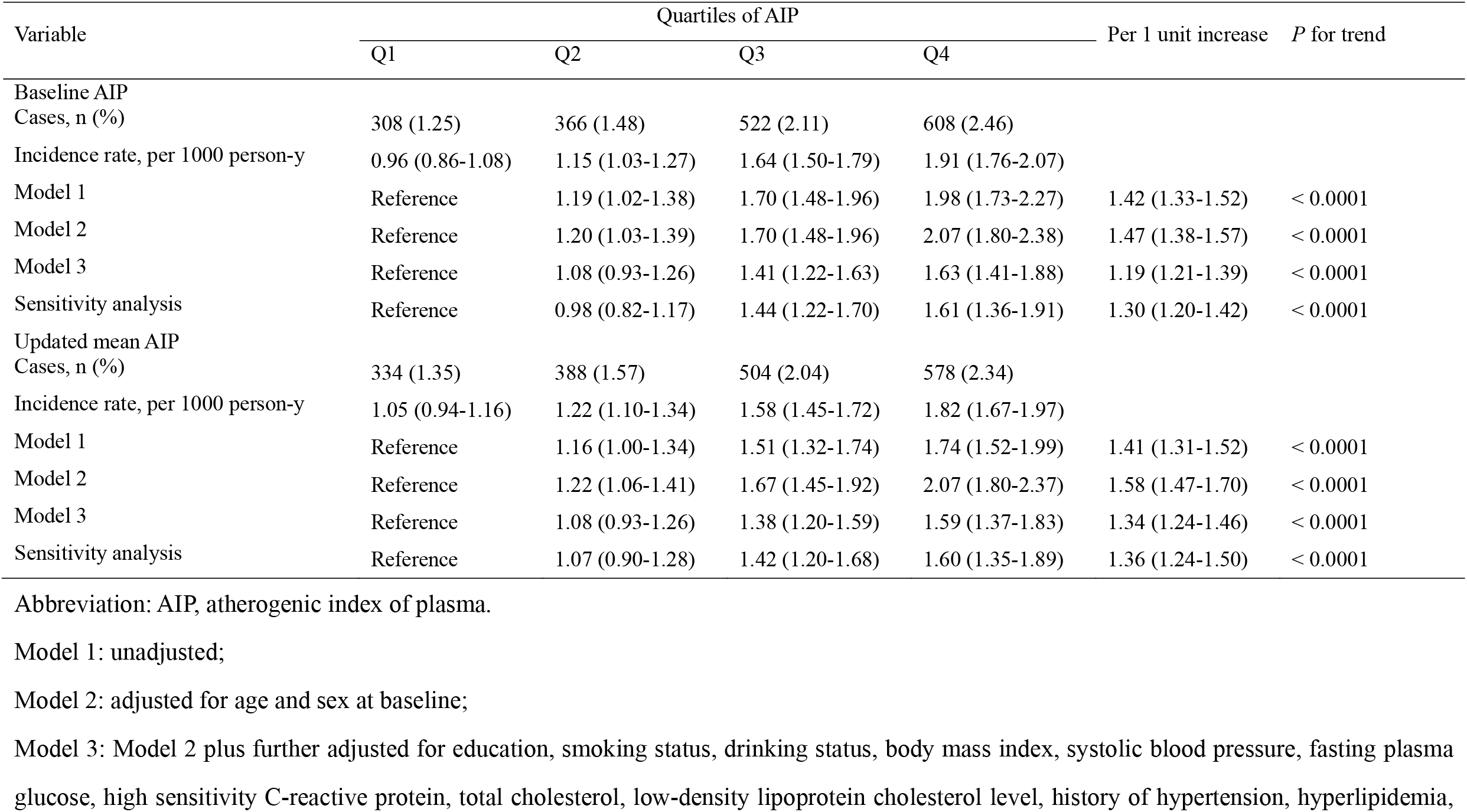
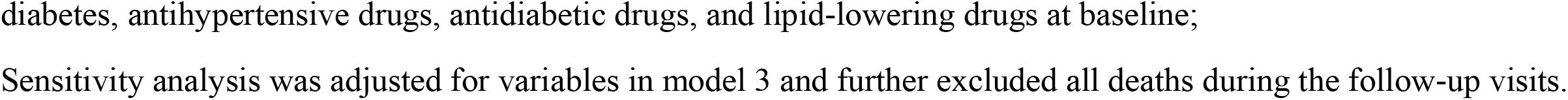
Association between quartiles of AIP with the risk of Myocardial Infarction

| Variable | Quartiles of AIP |  |  |  | Per 1 unit increase | P for trend |
| --- | --- | --- | --- | --- | --- | --- |
|  | Q1 | Q2 | Q3 | Q4 |  |  |
| Baseline AIP Cases, n (%) | 308 (1.25) | 366 (1.48) | 522 (2.11) | 608 (2.46) |  |  |
| Incidence rate, per 1000 person-y | 0.96 (0.86-1.08) | 1.15 (1.03-1.27) | 1.64 (1.50-1.79) | 1.91 (1.76-2.07) |  |  |
| Model 1 | Reference | 1.19 (1.02-1.38) | 1.70 (1.48-1.96) | 1.98 (1.73-2.27) | 1.42 (1.33-1.52) | < 0.0001 |
| Model 2 | Reference | 1.20 (1.03-1.39) | 1.70 (1.48-1.96) | 2.07 (1.80-2.38) | 1.47 (1.38-1.57) | < 0.0001 |
| Model 3 | Reference | 1.08 (0.93-1.26) | 1.41 (1.22-1.63) | 1.63 (1.41-1.88) | 1.19 (1.21-1.39) | < 0.0001 |
| Sensitivity analysis | Reference | 0.98 (0.82-1.17) | 1.44 (1.22-1.70) | 1.61 (1.36-1.91) | 1.30 (1.20-1.42) | < 0.0001 |
| Updated mean AIP Cases, n (%) | 334 (1.35) | 388 (1.57) | 504 (2.04) | 578 (2.34) |  |  |
| Incidence rate, per 1000 person-y | 1.05 (0.94-1.16) | 1.22 (1.10-1.34) | 1.58 (1.45-1.72) | 1.82 (1.67-1.97) |  |  |
| Model 1 | Reference | 1.16 (1.00-1.34) | 1.51 (1.32-1.74) | 1.74 (1.52-1.99) | 1.41 (1.31-1.52) | < 0.0001 |
| Model 2 | Reference | 1.22 (1.06-1.41) | 1.67 (1.45-1.92) | 2.07 (1.80-2.37) | 1.58 (1.47-1.70) | < 0.0001 |
| Model 3 | Reference | 1.08 (0.93-1.26) | 1.38 (1.20-1.59) | 1.59 (1.37-1.83) | 1.34 (1.24-1.46) | < 0.0001 |
| Sensitivity analysis | Reference | 1.07 (0.90-1.28) | 1.42 (1.20-1.68) | 1.60 (1.35-1.89) | 1.36 (1.24-1.50) | < 0.0001 |

Abbreviation: AIP, atherogenic index of plasma.
Model 1: unadjusted;
Model 2: adjusted for age and sex at baseline;
Model 3: Model 2 plus further adjusted for education, smoking status, drinking status, body mass index, systolic blood pressure, fasting plasma glucose, high sensitivity C-reactive protein, total cholesterol, low-density lipoprotein cholesterol level, history of hypertension, hyperlipidemia,
diabetes, antihypertensive drugs, antidiabetic drugs, and lipid-lowering drugs at baseline;
Sensitivity analysis was adjusted for variables in model 3 and further excluded all deaths during the follow-up visits.

Compared with quartile 1 group, the risk of MI increased significantly even after adjusting for potential confounding factors, the fully adjusted HRs (model 3) were 1.08 (95% CI, 0.93-1.26), 1.41 (95% CI, 1.22-1.63), and 1.63 (95% CI, 1.41-1.88) for quartiles 2, 3 and 4, respectively, versus quartile 1 of the baseline AIP (*P* for trend < 0.0001; Table 2). The association between the updated mean AIP with the risk of MI remained significantly, the adjusted HR in quartile 4 versus quartile 1 was 1.59 (95% CI, 1.37-1.83, *P* for trend <0.0001; Table 2). With death being regarded as a competing risk event, the result of sensitivity analysis showed the same association.

When AIP was treated as a continuous variable, a per 1 unit increase of the baseline AIP was associated with a 30% higher risk of MI (HR, 1.30; 95% CI, 1.20-1.42; Table 2), and a per 1 unit increase of the updated mean AIP was associated with a 36% higher risk of MI (HR, 1.36; 95% CI, 1.24-1.50; Table 2). After adjusting for all the potential confounding factors, the restricted spline curves for the associations between baseline AIP or the updated mean AIP and the risk of MI showed a *J*-shaped (Fig. 2).

**Figure. 2.**
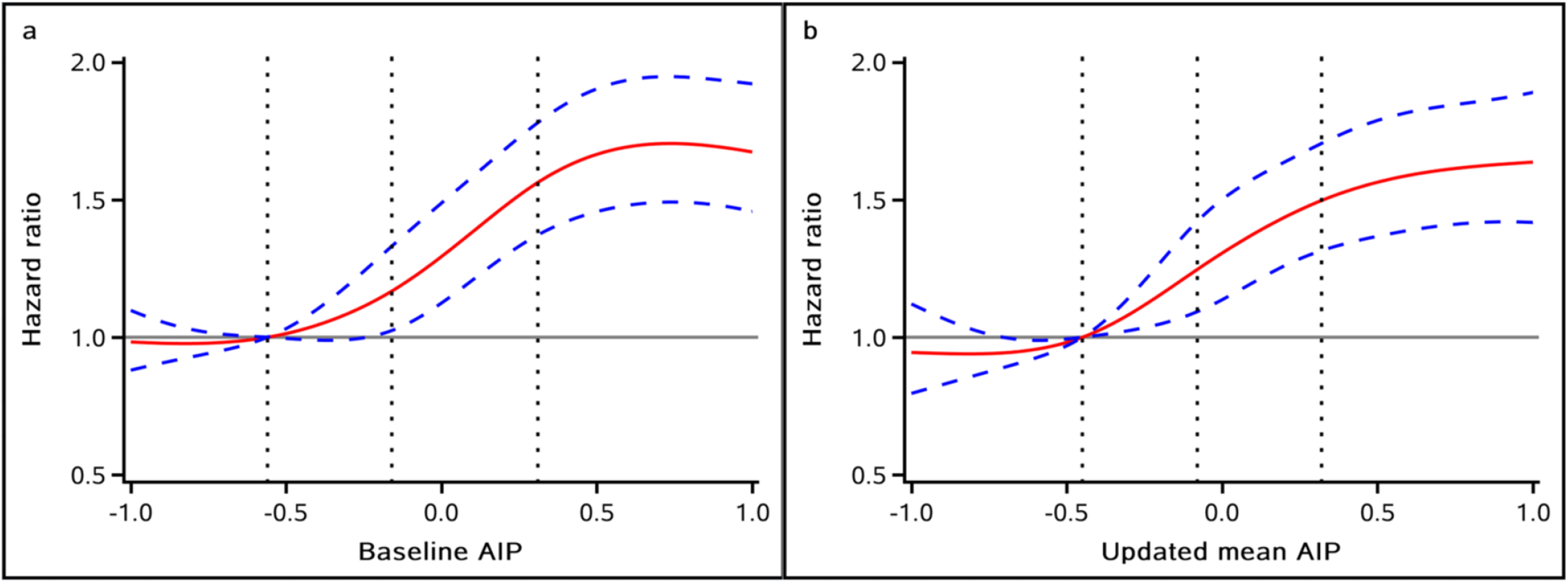
Multivariable-adjusted hazard ratios for MI based on restricted cubic spines with 5 knots at 5th, 25th, 50th, 75th, and 95th percentiles of a baseline AIP and b updated mean AIP. Abbreviation: AIP, atherogenic index of plasma. Red lines represent references for hazard ratios, and blue lines represent 95% confidence interval. Adjusted for age, sex, education, smoking status, drinking status, body mass index, systolic blood pressure, fasting plasma glucose, high sensitivity C-reactive protein, total cholesterol, low-density lipoprotein cholesterol level, history of hypertension, hyperlipidemia, diabetes, antihypertensive drugs, antidiabetic drugs, and lipid-lowering drugs at baseline.

A total of 56 012 eligible participants were included for the analysis between the number of visits with a high AIP and the risk of MI. The area under the ROC curve of AIP in case of incident MI was 0.60 (95% CI, 0.58-0.62), and the best cutoff point of AIP was 0.09 (Figure S1). The more times of visits with a high AIP, the risk of MI increased significantly. The fully adjusted HRs (model 3) were 1.63 (95% CI, 1.32-2.01), 1.93 (95% CI, 1.55-2.41), and 1.94 (95% CI, 1.55-2.42) for those with a high AIP one, two, three times, compared to those without high AIP at all the three visits. When death was regarded as a competing risk event, the sensitivity analysis showed similar association (Table 3).

**Table 3.** Association between number of visits with a high AIP with the risk of Myocardial Infarction

| Variable | Number of visits with a high AIP * |  |  |  | *P* for trend |
| --- | --- | --- | --- | --- | --- |
|  | 0 | 1 | 2 | 3 |  |
| Cases, n (%) | 178 (0.74) | 168 (1.29) | 160 (1.65) | 163 (1.74) |  |
| Incidence rate, per 1000 person-y | 0.57 (0.49-0.66) | 0.99 (0.85-1.16) | 1.28 (1.09-1.49) | 1.35 (1.16-1.57) |  |
| Model 1 | Reference | 1.80 (1.46-2.22) | 2.33 (1.88-2.89) | 2.48 (2.00-3.07) | < 0.0001 |
| Model 2 | Reference | 1.63 (1.32-2.01) | 1.93 (1.55-2.41) | 1.94 (1.55-2.42) | < 0.0001 |
| Model 3 | Reference | 1.63 (1.32-2.01) | 1.93 (1.55-2.41) | 1.94 (1.55-2.42) | < 0.0001 |
| Sensitivity analysis | Reference | 1.55 (1.22-1.96) | 1.86 (1.46-2.38) | 1.92 (1.50-2.45) | < 0.0001 |

Abbreviation: AIP, atherogenic index of plasma.
\* High AIP was defined as $AIP \geq -0.09$
Model 1: unadjusted;
Model 2: adjusted for age and sex at baseline;
Sensitivity analysis was adjusted for variables in model 3 and further excluded all deaths during the follow-up visits.

Subgroup analyses indicated that the higher risk of MI among participants with higher AIP level was consistent across relevant subgroups (*P* for interaction > 0.05 for all), including age (≤65 years vs. >65 years), sex (female vs. male), BMI (≤24 vs. ≥ 24 kg/m^2^), hyperlipidemia (no vs. yes), lipid-lowering drugs (no vs. yes), indicating no significant effect modification of the association between updated mean AIP and MI except BMI. In the BMI≤24 kg/m^2^ subgroup the fully adjusted HR was 1.78 (95% CI, 1.38-2.30), was higher than the BMI>24 kg/m^2^ subgroup (HR, 1.49; 95% CI, 1.25-1.79; Table 4). There were no significant effects between the baseline AIP and MI in subgroups (Table S2).

**Table 4.**
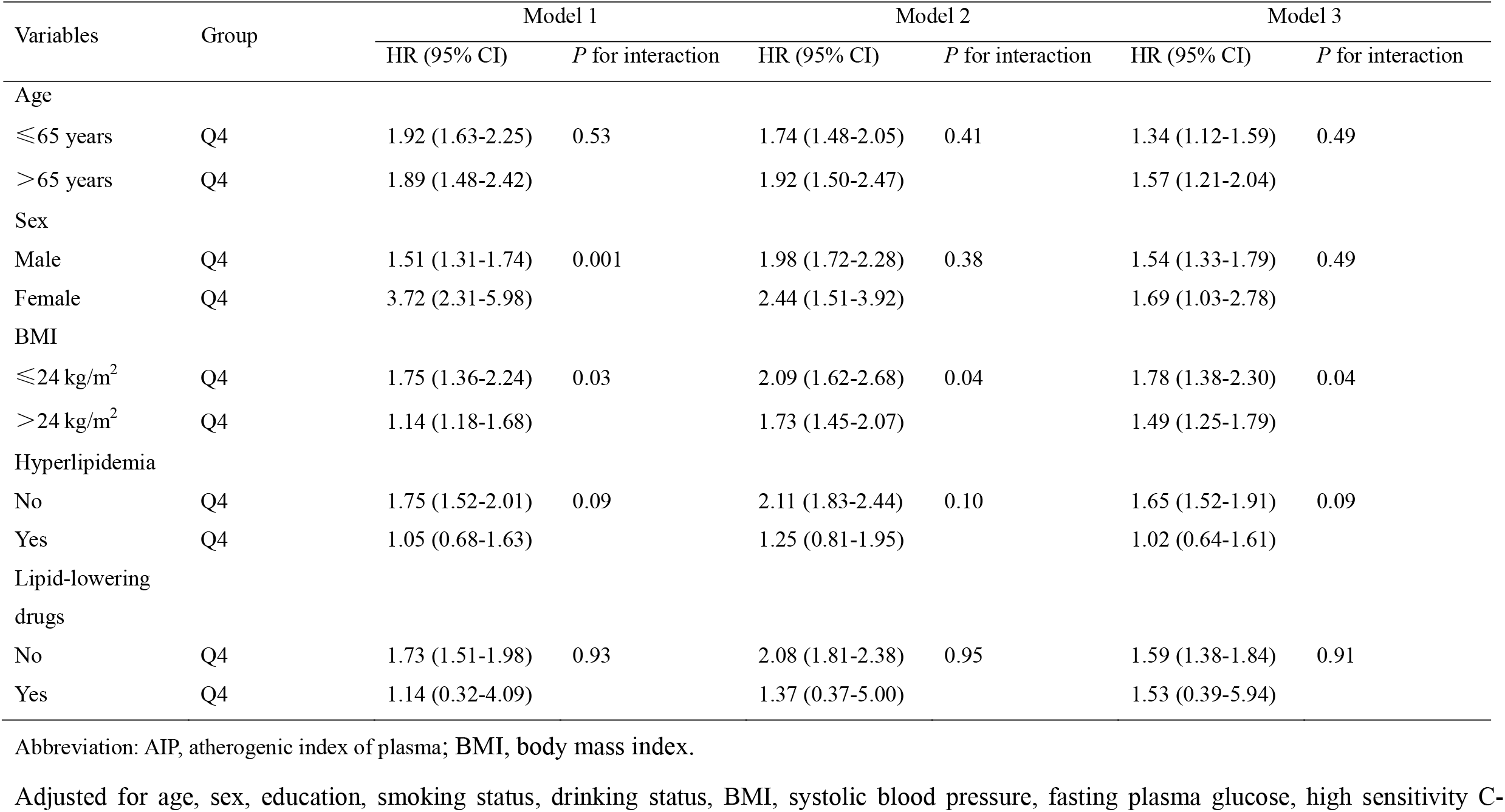
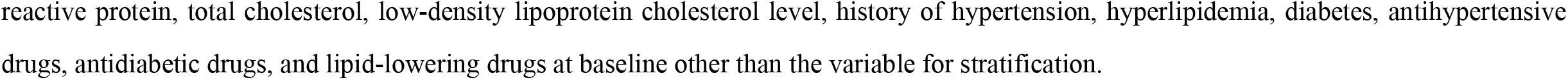
Association between updated mean AIP and the risk of Myocardial Infarction with stratification by baseline characteristics

| Variables | Group | Model 1 |  | Model 2 |  | Model 3 |  |
| --- | --- | --- | --- | --- | --- | --- | --- |
|  |  | HR (95% CI) | *P* for interaction | HR (95% CI) | *P* for interaction | HR (95% CI) | *P* for interaction |
| Age |  |  |  |  |  |  |  |
| ≤65 years | Q4 | 1.92 (1.63-2.25) | 0.53 | 1.74 (1.48-2.05) | 0.41 | 1.34 (1.12-1.59) | 0.49 |
| >65 years | Q4 | 1.89 (1.48-2.42) |  | 1.92 (1.50-2.47) |  | 1.57 (1.21-2.04) |  |
| Sex |  |  |  |  |  |  |  |
| Male | Q4 | 1.51 (1.31-1.74) | 0.001 | 1.98 (1.72-2.28) | 0.38 | 1.54 (1.33-1.79) | 0.49 |
| Female | Q4 | 3.72 (2.31-5.98) |  | 2.44 (1.51-3.92) |  | 1.69 (1.03-2.78) |  |
| BMI |  |  |  |  |  |  |  |
| ≤24 kg/m<sup>2</sup> | Q4 | 1.75 (1.36-2.24) | 0.03 | 2.09 (1.62-2.68) | 0.04 | 1.78 (1.38-2.30) | 0.04 |
| >24 kg/m<sup>2</sup> | Q4 | 1.14 (1.18-1.68) |  | 1.73 (1.45-2.07) |  | 1.49 (1.25-1.79) |  |
| Hyperlipidemia |  |  |  |  |  |  |  |
| No | Q4 | 1.75 (1.52-2.01) | 0.09 | 2.11 (1.83-2.44) | 0.10 | 1.65 (1.52-1.91) | 0.09 |
| Yes | Q4 | 1.05 (0.68-1.63) |  | 1.25 (0.81-1.95) |  | 1.02 (0.64-1.61) |  |
| Lipid-lowering drugs |  |  |  |  |  |  |  |
| No | Q4 | 1.73 (1.51-1.98) | 0.93 | 2.08 (1.81-2.38) | 0.95 | 1.59 (1.38-1.84) | 0.91 |
| Yes | Q4 | 1.14 (0.32-4.09) |  | 1.37 (0.37-5.00) |  | 1.53 (0.39-5.94) |  |

Abbreviation: AIP, atherogenic index of plasma; BMI, body mass index.
Adjusted for age, sex, education, smoking status, drinking status, BMI, systolic blood pressure, fasting plasma glucose, high sensitivity C-
reactive protein, total cholesterol, low-density lipoprotein cholesterol level, history of hypertension, hyperlipidemia, diabetes, antihypertensive drugs, antidiabetic drugs, and lipid-lowering drugs at baseline other than the variable for stratification.

## Data Availability

The data and SAS code used in this current study are available from the corresponding authors on a reasonable request.

## Discussion

This study of 98 861 individuals from the Kailuan Study found that both high baseline and long-term AIP were associated with increased risk of MI. Specifically, participants with the highest level of baseline and updated mean AIP, and those had three times of visits with a high AIP had the highest risk of MI. This novel finding indicated that risk of MI was better reflected by elevated levels of short-term or long-term AIP.

MI is still one of the top causes of death in China. As shown in various pathologic conditions, studies have shown that dyslipidemia was the most important risk factor of MI^20, 21^. Nonetheless, the great majority of previous studies have shown that the traditional single index of blood lipid as the evaluation of MI was still debated and not of high predictive value^22-24^.

The result of a cross-sectional study in healthy Korean adults showed the higher AIP level was an independent predictor of increased arterial stiffness^25^. A propensity score matching case-control study indicated that AIP might be a strong marker for predicting the risk of CVD in postmenopausal women^26^. A prospective study of 5538 non-diabetic CVD patients who had received percutaneous coronary intervention demonstrated that compared with tertile 1 of AIP, the risk of developing MACEs was 1.36-fold in the tertile 3 group^27^. A secondary analysis of the Action to Control Cardiovascular Risk in Diabetes (ACCORD) study showed AIP was an independent risk factor among the patients with type 2 diabetes mellitus AIP^28^. A recent registry study revealed that AIP could be associated with the outcome of dysfunction in patients with acute ischemic stroke, and this study also further evaluated the predictive values of blood lipid and found AIP was the best discriminating variable to predict poor outcomes, death, and disability^29^. In line with those past studies, our study of 98 861 participants with significant statistical power showed that the quartile 4 group of the baseline AIP had a 1.98-fold higher risk of MI compared with the quartile 1 group, and this significant association of a higher risk of MI in the quartile 4 group persisted after adjusting all potential covariates.

Previous studies focused on single time point to measuring AIP and the outcome at most, but ignored the longitudinal pattern of AIP during follow-up, and measurements of long-term exposure of AIP provide more reliable and robust results. In our study, we calculated the updated mean AIP and the number of visits with high AIP to determine the long-term AIP patterns of individuals. There have been some studies used these methods to explore the association between other parameters and the risk of CVD. The study from the Chronic Renal Insufficiency Cohort showed that time-updated SBP showed a stronger relation with chronic kidney disease than baseline SBP^30^. Another Kailuan study revealed that the participants with three visits with high triglyceride-glucose index had higher risk of developing MI than those without high triglyceride-glucose index^31^. Using the above methods, our study revealed that the risk of MI was highest in the quartile 4 group of the updated mean AIP and three visits with a high AIP. This result revealed that the high long-term AIP related to the occurrence risk of MI.

Subgroup analyses also indicated that the higher risk of MI among participants with higher baseline or updated mean AIP level. In the lower BMI subgroup, the risk of MI among participants with higher updated mean AIP level was higher. Stratified by BMI, a positive association between updated mean AIP and incident MI was more pronounced among individuals BMI≤24 kg/m^2^ than those BMI>24 kg/m^2^. Lower BMI tend to experience greater gains in AIP than higher. A previous study indicated that those individuals with normal weight but with metabolic abnormalities have a higher risk of CVD^32^. Our study revealed that both the elevated baseline and long-term AIP increased the risk of MI.

The strengths of the study include its prospective design, large community-based sample, long follow-up period, and the availability of repeated measurements of AIP. But this study also has several limitations. Firstly, this was an observational study, so we could not establish a causal association between AIP and the risk of MI, though we adjusted several confounding factors such as demographic characteristics, lifestyle, disease history and so on. There might be another included in the study that we could not control for, so our findings need to be confirmed in future studies. Secondly, since this study was based on the Northern Chinese coal miners, who were mainly males, our findings might not apply to other populations. However, on the other hand, the population in our study was quite homogeneous, which to some extent making our findings more reliable.

In conclusion, our study shows that an elevated AIP at baseline and in the long-term were independently associated with increased risk of MI. Our findings indicate that AIP may be useful for identifying individuals at high risk of developing MI, and emphasize the importance of monitoring AIP in the long term in clinical practice.

## Authors’ contributions

YZ and SW wrote the manuscript. YZ, AW, XT and JL researched the data. QX, XX, XZ, and SC researched the data and contributed to the discussion. AW and FL contributed to the discussion and reviewed/edited the manuscript. All authors read and approved the final manuscript.

## Acknowledgments

We thank all study participants, their relatives, the members of the survey teams at the 11 regional hospitals of the Kailuan Medical Group; and the project development and management teams at the Beijing Tiantan Hospital and the Kailuan Group.

## Ethics approval

The study was performed according to the guidelines of the Helsinki Declaration and was approved by the Ethics Committee of Kailuan Hospital (approval number: 2006e05) and Beijing Tiantan Hospital (approval number: 2010-014-01).

## Consent to participate

All the participants provided written informed consent.

## Consent for publication

Not applicable.

---

**Figure S1.**
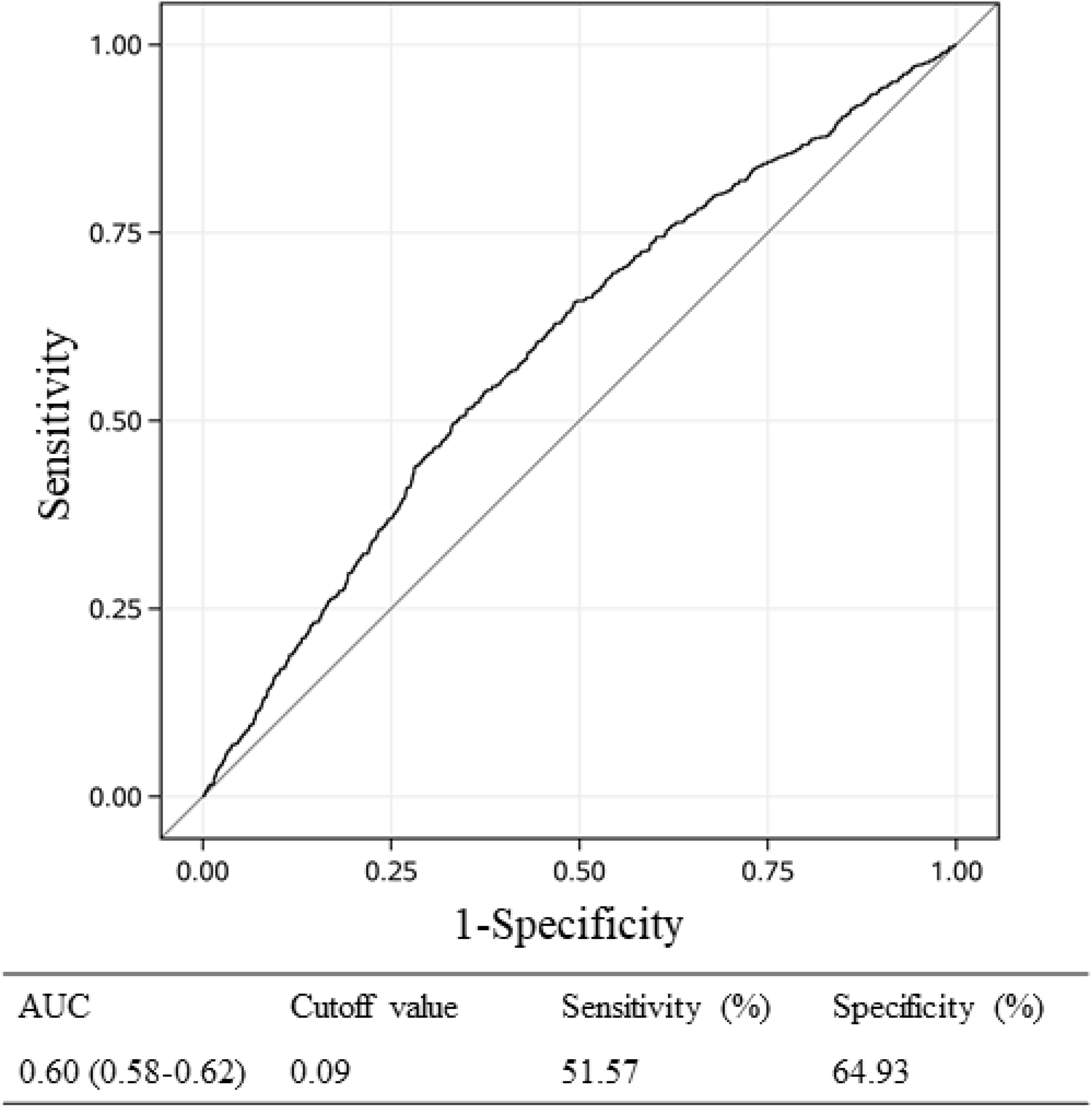
Receiver operative characteristics curve and cutoff value of AIP for incident MI. Abbreviation: AUC, area under curve; AIP, atherogenic index of plasma; MI, myocardial infarction.

**Table S1.**
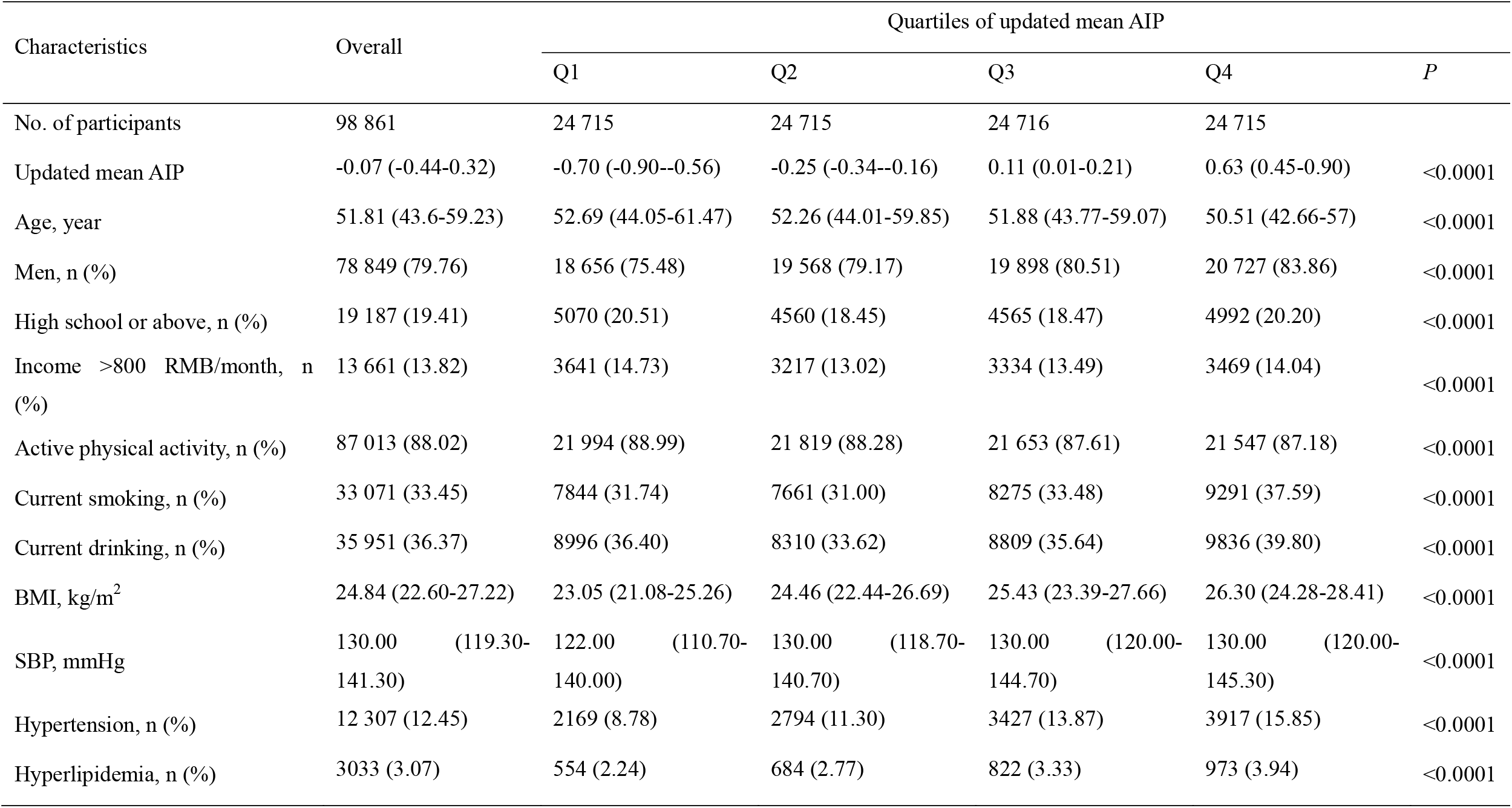
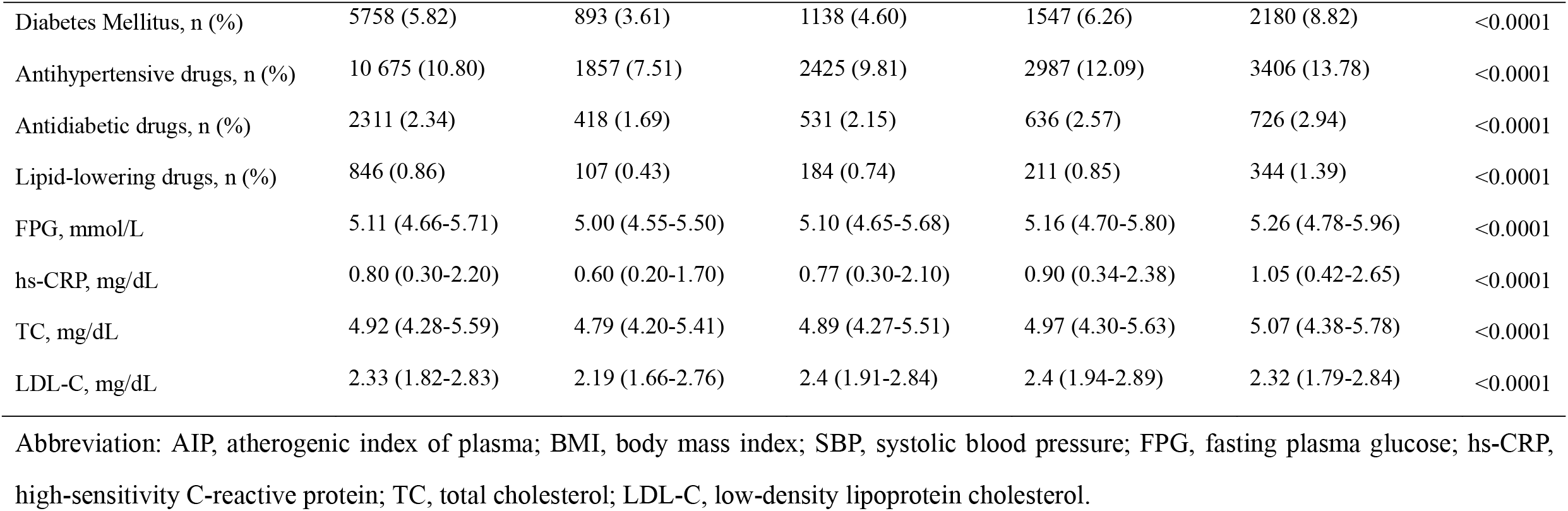
Baseline characteristics of participants according to quartiles of the updated mean AIP

**Table S2.**
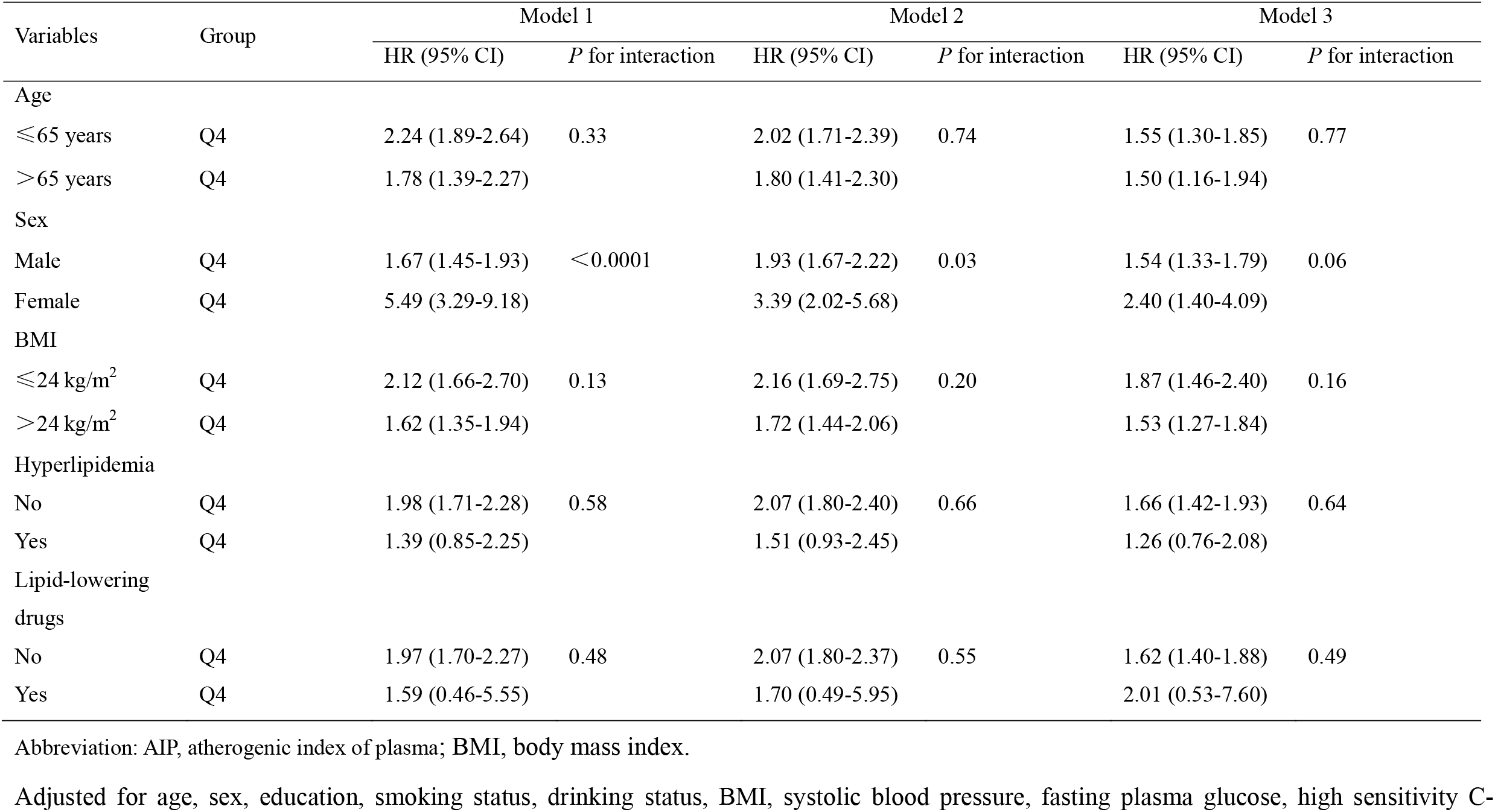
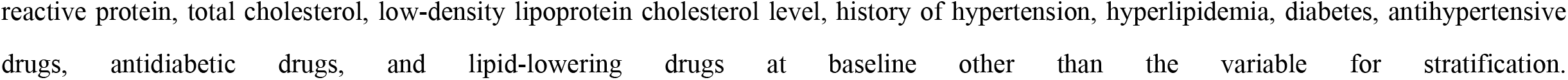
Association between AIP and the risk of Myocardial Infarction with stratification by baseline characteristics

| Variables | Group | Model 1 |  | Model 2 |  | Model 3 |  |
| --- | --- | --- | --- | --- | --- | --- | --- |
|  |  | HR (95% CI) | *P* for interaction | HR (95% CI) | *P* for interaction | HR (95% CI) | *P* for interaction |
| Age |  |  |  |  |  |  |  |
| ≤65 years | Q4 | 2.24 (1.89-2.64) | 0.33 | 2.02 (1.71-2.39) | 0.74 | 1.55 (1.30-1.85) | 0.77 |
| >65 years | Q4 | 1.78 (1.39-2.27) |  | 1.80 (1.41-2.30) |  | 1.50 (1.16-1.94) |  |
| Sex |  |  |  |  |  |  |  |
| Male | Q4 | 1.67 (1.45-1.93) | <0.0001 | 1.93 (1.67-2.22) | 0.03 | 1.54 (1.33-1.79) | 0.06 |
| Female | Q4 | 5.49 (3.29-9.18) |  | 3.39 (2.02-5.68) |  | 2.40 (1.40-4.09) |  |
| BMI |  |  |  |  |  |  |  |
| ≤24 kg/m<sup>2</sup> | Q4 | 2.12 (1.66-2.70) | 0.13 | 2.16 (1.69-2.75) | 0.20 | 1.87 (1.46-2.40) | 0.16 |
| >24 kg/m<sup>2</sup> | Q4 | 1.62 (1.35-1.94) |  | 1.72 (1.44-2.06) |  | 1.53 (1.27-1.84) |  |
| Hyperlipidemia |  |  |  |  |  |  |  |
| No | Q4 | 1.98 (1.71-2.28) | 0.58 | 2.07 (1.80-2.40) | 0.66 | 1.66 (1.42-1.93) | 0.64 |
| Yes | Q4 | 1.39 (0.85-2.25) |  | 1.51 (0.93-2.45) |  | 1.26 (0.76-2.08) |  |
| Lipid-lowering drugs |  |  |  |  |  |  |  |
| No | Q4 | 1.97 (1.70-2.27) | 0.48 | 2.07 (1.80-2.37) | 0.55 | 1.62 (1.40-1.88) | 0.49 |
| Yes | Q4 | 1.59 (0.46-5.55) |  | 1.70 (0.49-5.95) |  | 2.01 (0.53-7.60) |  |

Abbreviation: AIP, atherogenic index of plasma; BMI, body mass index.
Adjusted for age, sex, education, smoking status, drinking status, BMI, systolic blood pressure, fasting plasma glucose, high sensitivity C-
reactive protein, total cholesterol, low-density lipoprotein cholesterol level, history of hypertension, hyperlipidemia, diabetes, antihypertensive drugs, antidiabetic drugs, and lipid-lowering drugs at baseline other than the variable for stratification.

